# Setting Mental Health Research Priorities in Norfolk and Suffolk: A Stakeholder Consultation

**DOI:** 10.64898/2026.01.02.25343276

**Authors:** Sherifat Oduola, Sol Morrissey, Amy Zile, Jay Balaam, Craig Morgan, Jayati Das-Munshi, Niall Broomfield, Joni Holmes, Zarnie Khadjesari, Helen Parretti, Kristy Sanderson

## Abstract

**Introduction:** Mental health problems disproportionately affect marginalised communities rural, coastal, and socioeconomically disadvantaged communities. This is especially true for Norfolk and Suffolk, UK, where anxiety and depression are above national averages, suicide is the leading cause of death among those with mental ill-health, and access to care is poor. These communities are also underserved by research, leaving significant needs unmet.

**Aims:** This project aimed to establish mental health research priorities informed by the views of key stakeholders in Norfolk and Suffolk, including people with lived experience of mental ill-health, members of the public, clinicians, charities, and policymakers.

**Methods and analysis:** We conducted a mixed-methods research-priority-setting exercise involving an online survey (n=156, of whom 64.7% had lived experience) and two in-person prioritisation workshops with people with lived experience (n=10) and health and social care professionals (n=15). Following Delphi principles and guided by the James Lind Alliance approach, the survey asked participants to rate research priority statements across eight mental-health domains using a 3-point Likert scale (0 = low, 1 = moderate, 2 = high). Statements rated as high priority by ≥50% of the respondents were shortlisted for the workshops. During the two-day workshops, participants discussed the shortlisted statements in small groups before voting individually on those they considered most important. Scores were calculated separately for each workshop, then combined to produce a final ranked list of the Top 10 research priorities.

**Ethics:** The University of East Anglia’s Faculty of Medicine and Health Research Ethics Committee granted ethical approval (reference: ETH2324-2542).

**Results:** Of the 70 original priority statements, 40 met the threshold for inclusion in the prioritisation workshops. Participants in the two prioritisation workshops identified and agreed on the final Top 10 priorities, spanning youth mental health, physical–mental health integration, access to care, impacts of rural and coastal living, social and health inequalities, health promotion and prevention, and big-data solutions.

**Conclusions:** Stakeholders in this study identified local mental health needs and highlighted areas where research is urgently required. These priorities will inform future studies, support policymaking, and guide resource allocation to improve mental healthcare and reduce inequalities in rural and coastal communities.

**Strengths and limitations of this study:**

- This study brought together key stakeholders, including people with lived experience, mental health service providers, local authorities, policymakers, voluntary organisations, academics, and members of the public, to identify mental health research priorities specific to Norfolk and Suffolk populations.
- Our in-depth research priority-setting exercise identified the Top 10 research priority areas to inform future research and service planning.
- While the current study included family carers or parents of children with mental health difficulties, future mental health research priority-setting involving children is still needed.

## BACKGROUND

### Burdens of mental ill-health

Mental health problems are rising worldwide and disproportionately affect underserved communities, including rural, coastal, and socioeconomically disadvantaged groups [1]. In the UK, conditions such as anxiety, depression, schizophrenia, bipolar disorder, eating disorders, addictions, autism spectrum, and personality disorders cost the NHS over £118 billion annually [2], and the individual social and economic burdens are considerable [3, 4]. Several environmental, social, and psychological risk factors are linked to the onset of mental illness, including deprivation, social fragmentation, minority status, and childhood adversity [5–7], with significant regional disparities in incidence/prevalence, care, and outcomes [8, 9].

### Mental Health Challenges in Rural and Coastal Communities

Coastal communities face significant public health challenges in particular, including a high burden of physical and mental ill-health. They also have poor access to care, which is set to worsen without a vigorous and systematic approach to addressing them [10]. These challenges are mainly driven by socioeconomic stagnation and environmental factors, including deprivation, physical isolation, deindustrialisation, transient populations, and inward and outward migration of older and younger people [11, 12]. Additionally, rural communities are underserved by research and areas with the greatest mental health needs are under-represented or not engaged in research [13]. The paucity of UK research investigating patterns of access to care for, and long-term outcomes of mental illness among people living in rural communities may be an indication of the complexities of broader health service resource allocation and infrastructure, leading to significant barriers to accessing support in rural communities.

Norfolk and Suffolk, home to 1.6 million residents across rural, coastal, and urban areas, have above-average rates of anxiety and depression, high suicide risk, and poor access to services [14, 15]. Furthermore, mental health outcomes in coastal areas, including in our case Great Yarmouth and Lowestoft, are complicated by high levels of homelessness, alcohol use, and substance misuse [12, 16]. However, rural and coastal communities remain under-represented in research, and there are very few interventions available that address these complex factors alongside mental health support and prevention.

### Young People’s Mental Health

Globally, mental ill-health contributes to three of the top six causes of disability-adjusted life years among 10–24-year-olds, with suicide the third leading cause of death [17, 18]. Considering the potential negative impact of mental illness on adult outcomes, adolescence is a critical period for early detection, rapid access to support and treatment, and for mental health promotion. This is especially important in rural areas, where barriers like stigma, limited-service awareness, and confidentiality concerns hinder help-seeking [19, 20]

### Social Disadvantage and Mental Health

Mental health problems disproportionately affect socioeconomically disadvantaged groups, those in fragmented or deprived communities [21, 22], and those exposed to adversity [23]. Marginalised populations, including racially minoritised groups and those with severe mental illness, face compounded disadvantages through discrimination and poor access to care [24–26]. Yet our understanding of how social and environmental conditions shape mental health trajectories, recovery, and resilience remains limited, particularly outside urban settings.

### Premature Mortality and Multimorbidity

People with severe mental illness die 15–20 years earlier than the general population [27, 28], largely due to multimorbidity, including cardiovascular disease, diabetes, and stroke [29, 30]. How and why this occurs, and why this disproportionately affects people living in rural/coastal areas is poorly understood. While there have been calls for greater integration between mental and physical health (e.g., *No Health without Mental Health* [31] and *Supporting the physical health of people with severe mental illness* [32]), progress has been slow in narrowing the mortality gap.

### Rationale, Aims and Objectives

Addressing disparities in mental healthcare and research is a central priority of the World Health Organisation (WHO) strategy for strengthening health systems globally [33]. But, in general research on mental health is not conducted in areas with the highest levels of need. If we are to have a full understanding of both the determinants of mental ill health and how to respond effectively, we need to expand our geographical scope; a key step in this is understanding local priorities for research and intervention.

Furthermore, for research to generate meaningful impact, it must be developed and prioritised in partnership with key stakeholders, particularly those most affected by mental health challenges. Guided by a community-based participatory research approach [34], we sought to identify the mental health needs of communities across Norfolk and Suffolk, and to establish a shared set of research priorities through collaboration with stakeholders, local partners, and experts, laying the foundation for a long-term research programme and joint working. We addressed the following objectives:

1. Identify and bring together people with lived experience, mental health service providers, local authorities, policymakers, charities, educators, and researchers in Norfolk and Suffolk.
2. Assess the relevance of the available research priorities using the James Lind Alliance’s “overarching priority topics” for the health and care research toolkit to identify areas of need through consultation.
3. Work with stakeholders to produce a list of the Top 10 applied mental health research priorities in Norfolk and Suffolk to inform future research programmes.

## METHODS

### Study design and procedure

In this paper -funded by the National Institute of Health Research (NIHR207498) -we present findings from a priority setting exercise to understand local needs; what we found may resonate with similar places and how we conducted the study provides a template for others to apply. And if applied by others it will enable the accumulation of comparable data on variations and similarities in priorities and inform a new generation of place based mental health research. We have published our study protocol in [1] and the study is reported in accordance with the ACCORD Research Checklist [35].

Our study drew on the principles of Delphi methodology and was guided by James Lind Alliance (JLA) framework. The JLA is an interpersonal framework to build consensus and is the mostly used approach in setting health research priorities [36]. We chose the JLA for its structured approach to inclusivity, transparency, and the direct alignment of research goals with the lived experiences of patients and clinicians. We identified and adapted existing mental health research priorities from the JLA “overarching priority topics” for the health and care research toolkit [37] to establish their relevance to populations in rural/deprived areas of Norfolk and Suffolk and whether there were unanswered questions/topics that, if examined by research, could make a real difference to people’s lives. Figure 1 summarises the roadmap of the phases of our research priority setting exercise.

**Figure 1:**
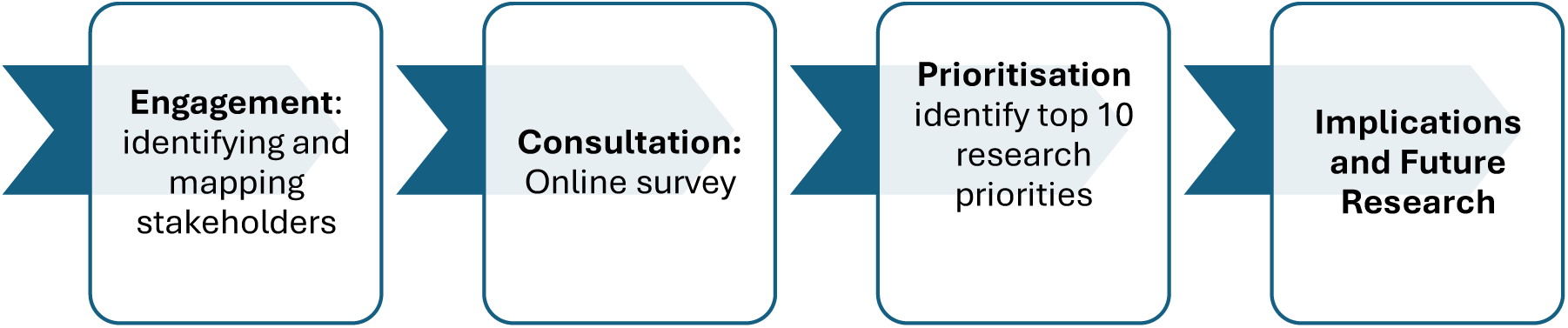
Roadmap of research priority-setting exercise

#### Study settings, participants and eligibility

Stakeholders aged ≥16 years and fluent in the English language were eligible to participate. This included people with lived experience of mental ill-health, carers or friends of someone with a mental ill-health, mental health service providers, other health and social care professionals, local authorities, policymakers, charities, youth groups, educators, community groups, and researchers. All participants were sampled from communities and organisations across Norfolk and Suffolk; everyone meeting the above criteria was considered eligible to participate in the study, and no further exclusion criteria were applied.

### Patient and Public Involvement and Engagement (PPIE)

Two advisory groups supported the study from the outset. **The Lived Experience Advisory Group (LEAG)**: The LEAG, led by our PPI lead (a peer researcher), comprised five members with lived experience of mental health difficulties or of caring for someone with a mental health condition. The LEAG contributed to and informed the survey design, prioritisation workshops, and interpretation of findings. Meetings were held in-person and online, in a group or 1:1 setting (depending on the member’s preferences), between June 2024 and May 2025. Members were reimbursed for their time at NIHR recommended rates.

**Steering Group (SG):** comprised of health and social care professionals. Members were recruited via personal/professional networks and included clinicians, local authority public health experts, voluntary organisation representatives, and policy makers. The SG met three times (2 online and 1 in-person) between June 2024-May 2025 and provided insights into mental health needs and care delivery in Norfolk and Suffolk. Three SG members piloted the survey questionnaire ahead of recruitment. Their feedback (i.e. re-structuring aspects of the questionnaire for clarity) was incorporated into the final version.

### Ethics

This study was approved by the University of East Anglia’s Faculty of Medicine and Health Research Ethics Committee (reference: ETH2324-2542). Participants in the online survey had a chance to win one of 20 £25 gift vouchers as a thank-you for completing the questionnaire. Prioritisation workshop participants were offered a £25 gift voucher each as reimbursement for their time, and their travel expenses were covered. Informed consent was obtained from all participants prior to data collection commencing.

#### Phase 1 -Identifying stakeholders (study objective 1)

We identified stakeholders through research databases, professional networks, online searches, and partner organisations, then mapped into six expert pools: (1) people with lived experience/carers/public; (2) health and social care professionals; (3) third sector/community groups; (4) researchers; (5) local authorities, schools, employers; and (6) policymakers.

#### Phase 2 – Consultation- online survey (study objective 2)

We co-designed an online questionnaire with the SG and LEAG to gather stakeholders’ views on mental health research priorities. From the JLA “overarching priority” topics for the health and care research toolkit [37], we created a list of research priority statements across eight domains. These were 1. Children and young people’s mental health, 2. The link between physical and mental health, 3. The impact of rural and coastal living on mental health, 4. Access to mental healthcare, 5. Migration and mental health, 6. Social and health inequalities, 7. Mental health promotion and prevention and 8. Big Data Solutions. Free-text fields were included in the survey questionnaire for participants to list up to three additional priorities not included in the questionnaire. The survey questionnaire is available in [1].

##### Data collection

Stakeholders from the Phase 1 expert pools were invited to participate in a three-step data collection process. First, participants rated the importance of research priority statements on a 3-point Likert scale (0 = low, 1 = moderate, 2 = high), as recommended by JLA. Second, they listed up to three additional mental health research priorities in free-text responses. Third, the highest-rated statements (i.e. ≥50% rated as high-priority) were shortlisted for the Phase 3 prioritisation workshops.

Sociodemographic (age, sex, gender, ethnicity, employment, education) and geographical (rural, urban, coastal) characteristics were collected. Respondents with lived experience of mental ill-health provided details of diagnosis, illness duration, and access to treatment. Data were collected between September 2024 and January 2025 via partner organisations and networks that served as gatekeepers. We also promoted the survey link and QR code via social media, our websites, newsletters, and posters. Informed consent was obtained online prior to participation.

###### Sample size

The JLA does not recommend a minimum or maximum number of responses [38]. However, we aimed for 165 responses, which is an average response rate based on previous JLA priority-setting exercises with rapid approaches [39].

#### Phase 3 – Prioritisation workshops (study objective 3)

Our stakeholder research prioritisation exercise followed JLA principles through two workshops at the University of East Anglia in March 2025. Separate sessions were held for (1) service users/public, i.e. lived experience stakeholder group, and (2) health and social care professionals, i.e. professional group, aiming to review Phase 2 findings and agree on the Top 10 mental health research priorities to guide future research. There were thirty attendees: lived experience (n=10), professionals (n=15), and facilitators, including the PPI lead (n=5). Participants were recruited via our professional networks, partner organisations, websites, newsletters, posters, and those who had registered an interest in Phase 2. Written consent and brief demographic details were collected before participation. Members of the research team documented discussions and meeting notes to capture decision-making and participants’ views.

##### Data collection

Data collection was undertaken over two days with separate workshops and processes with two stakeholder groups: 1. people with lived experience of mental ill-health or of caring for someone with a mental health condition (referred to as the ‘lived experience group’), and 2. health and social care professionals (referred to as the ‘professional group’).

**Day 1:**

Ten people with lived experience of mental ill-health (including LEAG members) attended with the research team.

1. The session began with an introduction and a “get to know you” exercise, where the participants and the wider study team shared their motivation for taking part in the study. The chief investigator presented the study overview and the shortlisted priority statements from the survey; participants had the opportunity to ask questions.
2. Participants were divided into two groups, where they reviewed and discussed the shortlisted priority statements. Two researchers facilitated the group discussions. Participants voted on the shortlisted priority statements individually (high = 2, moderate = 1, low = 0).
3. Scores were analysed in real time (i.e. during the break) by two researchers using MS Excel.
4. The scores were presented to the whole group, and participants ranked the statements (1-10) to determine the final top 10 mental health research priorities.

**Day 2:**

Fifteen health and social care professionals representing statutory and voluntary organisations across Norfolk and Suffolk. The same format and steps 1-4 as described for Day 1 were followed. However, to ensure that lived experience and public view remained central to the priority-setting exercise, the professional group received the ranking scores and feedback from Day 1 and had the opportunity to refine or revise their responses. This adaptation has been used in previous studies, which show that providing feedback on patient scores to healthcare professionals results in a broader set of consensus items that better reflect patients’ priorities [40, 41].

##### Data analysis

Descriptive statistics were used to analyse the survey and prioritisation data. Content analysis was used to analyse the free-text data. Quantitative data were analysed using Microsoft Excel and in RStudio 4.3.1 [42]. The triangulation method was used to integrate information from the survey and free text. Triangulation is a recognised approach to achieving and maintaining consistency, validity, and rigour in mixed-methods research [43]. Eight participants were removed from the survey data analyses because they were neither residents nor worked in Norfolk, Suffolk, or East Anglia. Apart from the sample characteristics reported in Table 1, all other analyses were conducted with data from participants who lived or worked in the study area. Thematic analysis was used to summarise meeting notes from the 2-day workshops.

**Table 1.** Sample characteristics.

| Characteristics | N=156 (%) |
| --- | --- |
| **Sex (female)** | 119 (76.3) |
| **Gender** |  |
| Woman | 107 (68.6) |
| Man | 36 (22.4) |
| Transgender | 4 (2.6) |
| Non-binary | 3 (1.9) |
| Prefer not to say | 6 (3.8) |
| **Age group** |  |
| 16-19 | 33 (24.3) |
| 20-29 | 29 (21.3) |
| 30-39 | 22 (16.2) |
| 40-49 | 19 (14.0) |
| 50-59 | 31 (22.8) |
| 60-69 | 16 (11.8) |
| 70-79 | 6 (4.4) |
| 80-89 | 0 (0) |
| **Ethnicity** |  |
| White British (English/ Welsh/ Scottish/ Northern Irish) | 121 (77.6) |
| White non-British | 15 (9.6) |
| Mixed ethnic background | 7 (4.5) |
| Black ethnic group | 8 (5.1) |
| Indian | 2 (1.3) |
| Any other ethnic background | 1 (0.6) |
| Prefer not to say | 2 (1.3) |
| **Country of birth** |  |
| UK | 128 (90.1) |
| Non-UK | 14 (9.9) |
| **Mental health diagnosis** |  |
| Diagnosis status | 75 (48.1) |
| Diagnosis from UK | 74 (98.7) |
| Diagnosis from other country | 1 (1.3) |
| Duration of diagnosis | 11.5 (SD: xx) years |
| Receiving treatment | 60 (80.0) |
| **Employment status** (currently employed) | 101 (65.2) |
| **Stakeholder group\*** |  |
| People with lived mental health experience | 101 (64.7) |
| Carer (family or friend) | 65 (41.7) |
| Mental health professional | 29 (18.6) |
| Representative of a voluntary sector/ charity organization | 19 (12.2) |
| Other (Policymaker/researcher/educator) | 15 (9.6) |
| Member of the public interested in MH | 40 (25.6) |
| **Live or work in Norfolk, Suffolk or East Anglia** |  |
| Yes | 148 (94.9) |
| No | 8 (5.1) |
| **Area of Residence** |  |
| Norfolk | 92 (59.0) |
| Suffolk | 55 (35.3) |
| Other | 1 (0.7) |
| **Moved to East Anglia within 10 years** |  |
| No | 116 (74.8) |
| Yes | 40 (25.2) |
| **Moved from (if moved)** |  |
| Within the UK | 31 (83.8) |
| Outside the UK | 5 (13.5) |
| Prefer not to say | 1 (2.7) |
| **Geographical setting** |  |
| Coastal | 4 (2.6) |
| Hamlet/village | 15 (9.8) |
| Rural town and fringe | 51 (33.3) |
| Urban | 83 (54.2) |
| **Highest educational attainment** |  |
| No formal qualifications | 1 (0.6) |
| GCSE/ O' Level/ CSE/ National Qualifications/ Standard Grades | 31 (20.1) |
| Tertiary (e.g. A 'level) | 8 (5.2) |
| Higher (e.g. Diploma, HND) | 21 (13.6) |
| University (degree, postgraduate, PhD) | 79 (51.3) |
| Still studying | 13 (8.4) |
| I prefer not to say | 1 (0.6) |

\*Participants could identify as more than one group

## RESULTS

### Phase 1: Stakeholder mapping

Through the stakeholder mapping exercise, we identified 115 statutory and voluntary, community and social enterprise organisations that support people with mental health difficulties across Norfolk and Suffolk.

### Phase 2: Online Survey

Table 1 summarises the participants’ sociodemographic characteristics. Of the one hundred and fifty-six participants who completed the survey, the majority (n=101, 64.7%) identified as people with lived experience of mental ill-health, with a mean 11.5 (SD: 3.6) years duration of mental health diagnosis. Across the sample, there were more women (n=119, 76.3%), mostly of White British ethnic background (n=121, 77.6%), employed (n=101, 64.7%), and educated to university level (n=79, 51.3%). Geographically, most participants lived or worked in urban areas, with 33.3% from rural areas. Fewer (3%) participants were from coastal areas. A quarter (n=40) of the sample had moved to Norfolk, Suffolk and the East Anglia region in the last 10 years, of whom the majority (n=31, 83.8%) moved from within the UK.

Table 2 summarises the responses to the priority ratings by domains. All the domains were rated as high research priority, scoring 50% or higher, except migration and mental health, which scored 48.1%. Supplementary Table S1 presents the summary of responses to the priority statements across the key domains.

**Table 2:** Responses to research priority statements by domains (n=148)

| **Domain** | **High Priority** | **Medium Priority** | **Low Priority** | **Average score (SD)** |
| --- | --- | --- | --- | --- |
| Children and Young people's mental health (**n, %**) | 85 (58.1) | 49 (33.4) | 12 (8.5) | 1.5 (0.6) |
| The link between physical and mental health (**n, %**) | 80 (55.0) | 56 (38.3) | 10 (6.7) | 1.5 (0.6) |
| Access to mental healthcare (**n, %**) | 95 (65.1) | 43 (29.1) | 8 (5.8) | 1.6 (0.6) |
| The impact of rural and coastal living on mental health (**n, %**) | 72 (50.0) | 51 (35.3) | 22 (14.7) | 1.3 (0.7) |
| Migration and mental health (**n, %**) | 69 (48.1) | 56 (39.4) | 18 (12.4) | 1.4 (0.7) |
| The impact of social and health inequalities on mental health (**n, %**) | 89 (62.8) | 41 (28.8) | 12 (8.3) | 1.5 (0.6) |
| Mental health promotion and prevention (**n, %**) | 86 (60.9) | 45 (31.9) | 10 (7.2) | 1.5 (0.6) |
| Big data solutions for improving mental health outcomes (**n, %**) | 90 (62.0) | 48 (33.4) | 7 (4.6) | 1.6 (0.6) |

Table 3 presents the shortlisted priority statements (n=40), i.e. rated as high priority (scoring ≥50%).

**Table 3.** Shortlisted research priority statements.

| Domain | Priority Statement | High priority | Medium priority | Low priority | Total score: |
| --- | --- | --- | --- | --- | --- |
| Children and Young people's mental health | Involving young people in making decisions about their mental health treatment | 19 (73.1) | 7 (26.9) | 0 (0) | 45 |
|  | Child and Adolescent Mental Health Services (CAMHS), education providers, and health and social care departments work together more effectively to improve the mental health outcomes of children and young people | 19 (73.1) | 7 (26.9) | 0 (0) | 45 |
|  | Supporting young people on Child and Adolescent Mental Health Services (CAMHS) waiting lists to prevent further deterioration of their mental health | 19 (73.1) | 7 (26.9) | 0 (0) | 45 |
|  | Early intervention strategies for supporting children and young people in improving their mental resilience | 18 (69.2) | 7 (26.9) | 1 (3.8) | 43 |
|  | Training teachers and other staff in schools and colleges to detect early signs of mental health difficulties in children and young people | 17 (65.4) | 8 (30.8) | 1 (3.8) | 42 |
|  | Increase availability of effective culturally appropriate services in children and young people's mental health, particularly for ethnic minority groups | 16 (61.5) | 7 (26.9) | 3 (11.5) | 39 |
| The link between physical and mental health | Identifying the most effective and acceptable strategies for preventing multiple physical and mental health conditions in later life | 17 (65.4) | 9 (34.6) | 0 (0) | 43 |
|  | Treating mental and physical health together | 16 (61.5) | 9 (34.6) | 1 (3.8) | 41 |
|  | Identifying interventions that are most effective in improving sleep in people with mental health difficulties | 10 (38.5) | 16 (61.5) | 0 (0) | 36 |
|  | Improving physical health checks in people living with mental health difficulties | 11 (42.3) | 14 (53.8) | 1 (3.8) | 36 |
| The impact of rural and coastal living on mental health | Increasing access to psychological/emotional support for family carers/informal carers of people with mental health difficulties | 18 (69.2) | 8 (30.8) | 0 (0) | 44 |
|  | Improving access to mental health care for rural and coastal communities | 19 (73.1) | 5 (19.2) | 2 (7.7) | 43 |
|  | Improving access to mental health care for all age groups | 17 (65.4) | 9 (34.6) | 0 (0) | 43 |
|  | Improving access to psychological therapy-style services for children and adolescents | 18 (69.2) | 7 (26.9) | 1 (3.8) | 43 |
|  | Integrating the work of mental health charities with NHS services to improve access to care | 18 (69.2) | 7 (26.9) | 1 (3.8) | 43 |
|  | Addressing the barriers to accessing care and treatment for mental health difficulties in rural, remote and coastal areas | 18 (69.2) | 7 (26.9) | 1 (3.8) | 43 |
|  | Better awareness of where to seek help for mental health difficulties | 14 (53.8) | 10 (38.5) | 2 (7.7) | 38 |
|  | Understanding which models of care increase access and support decision-making for people with mental health difficulties | 12 (46.2) | 13 (50) | 1 (3.8) | 37 |
| Access to mental healthcare | More collaboration between health, social care, and voluntary sectors to deliver mental health care to people in rural areas | 19 (73.1) | 7 (26.9) | 0 (0) | 45 |
|  | Understanding the impacts of social isolation on physical and mental health | 16 (61.5) | 10 (38.5) | 0 (0) | 42 |
|  | Increase social and economic opportunities to enhance the mental health and wellbeing for young people and adults living in rural and coastal communities. | 17 (65.4) | 8 (30.8) | 1 (3.8) | 42 |
|  | Mental health team bases offering a single service that provides mental health support in rural/coastal areas | 16 (61.5) | 9 (34.6) | 1 (3.8) | 41 |
|  | Understanding the impact of rural/coastal living on recovery | 15 (57.7) | 10 (38.5) | 1 (3.8) | 40 |
|  | GP surgeries offering a single service that provides mental health support in rural/coastal areas | 14 (53.8) | 11 (42.3) | 1 (3.8) | 39 |
| Impact of social and health inequalities on mental health | Improving prevention efforts for suicide and self-harm in other jobs with higher risk of suicide | 19 (73.1) | 7 (26.9) | 0 (0) | 45 |
|  | Improving prevention efforts for suicide and self-harm in young people | 18 (69.2) | 8 (30.8) | 0 (0) | 44 |
|  | Understanding how effective community-based interventions are in improving mental health in deprived areas | 17 (65.4) | 8 (30.8) | 1 (3.8) | 42 |
|  | Identify mental health policy/interventions that could target key stages of life (over the life course) that could prevent poor mental health outcomes | 15 (57.7) | 10 (38.5) | 1 (3.8) | 40 |
|  | Develop culturally sensitive care for racial and ethnic minoritised groups | 15 (57.7) | 9 (34.6) | 2 (7.7) | 39 |
|  | Develop public health interventions to ensure no one is 'left behind' | 13 (50) | 12 (46.2) | 1 (3.8) | 38 |
|  | Improving prevention efforts for suicide and self-harm in farmers and agricultural communities | 13 (50) | 9 (34.6) | 3 (11.5) | 35 |
|  | Interventions for reducing involuntary hospital admissions among black and ethnic minoritised groups | 12 (46.2) | 10 (38.5) | 4 | 34 |
| Mental health promotion and prevention | Support for carers of older people with multiple conditions to maintain their psychological well-being | 17 (65.4) | 8 (30.8) | 1 (3.8) | 42 |
|  | Supporting parents or family members in identifying early signs of mental health difficulties in children and young people | 16 (61.5) | 8 (30.8) | 1 (3.8) | 40 |
|  | Early intervention to prevent mental health problems in people with disabilities. | 14 (53.8) | 12 (46.2) | 0 (0) | 40 |
|  | Support to find and retain employment among adults with mental health difficulties | 13 (50) | 13 (50) | 0 (0) | 39 |
|  | Empowering patients, relatives, and carers be to have a greater say about their choices regarding mental health and mental healthcare | 14 (53.8) | 11 (42.3) | 1 (3.8) | 39 |
|  | Understanding how mental health difficulties impact employment | 12 (46.2) | 13 (50) | 1 (3.8) | 37 |
| Big data solutions for improving mental health outcomes | Using de-identified data (e.g. health, education, local authority records) to examine mental health outcomes. | 15 (57.7) | 9 (34.6) | 2 (7.7) | 39 |
|  | Using de-identified data (e.g. health, education, local authority records) to identify risk factors of mental illness. | 15 (57.7) | 8 (30.8) | 3 (11.5) | 38 |

#### Qualitative free-text data

Fifty-eight participants completed the free-text fields indicating their own research priorities. All but 12 of the participants’ priorities aligned with and were mapped to the predefined statements (see Supplementary Table S2).

### Phase 3: Priority-setting workshops

Table 4 presents the characteristics of the participants who attended the two prioritisation workshops. A total of twenty-five participants (Lived experience group, n=10; Professional group, n=15) attended the two-day workshops. Most participants in both groups were women, aged 50-59 years, of White British ethnicity, and mostly lived or worked in Norfolk. Around a third of people in both groups had moved to the region in the last 10 years, and many had moved from within the UK. Participants from the professional group were mostly from rural areas (n=9, 60%) and worked within the voluntary sector (n=10, 66.7%). Of the participants in the lived experience group, two people identified as a family carer of someone with mental health difficulties,

**Table 4:** Prioritisation workshops participants demographic characteristics.

| Variable | Lived experience group<br>N = 10 (%) | Professionals group<br>N = 15 (%) |
| --- | --- | --- |
| **Age** |  |  |
| 20-29 | 2 (20) | 1 (6.7) |
| 30-39 | 1 (10) | 4 (26.7) |
| 40-49 | 1 (10) | 5 (33.3) |
| 50-59 | 3 (30) | 5 (33.3) |
| 60-69 | 2 (20) | 0 (0) |
| 70-79 | 1 (10) | 0 (0) |
| 80-89 | 0 (0) | 0 (0) |
| 90+ | 0 (0) | 0 (0) |
| **Sex** |  |  |
| Female | 6 (60) | 12 (80) |
| **Gender** |  |  |
| Man | 4 (40) | 3 (20) |
| Woman | 6 (60) | 12 (80) |
| Other | 0 (0) | 0 (0) |
| **Employment status** |  |  |
| Employed | 5 (50) | 15 (100) |
| Studying | 4 (40) | 0 (0) |
| **Ethnicity** |  |  |
| White British | 8 (80) | 12 (80) |
| White non-British | 2 (20) | 2 (13.3) |
| Black ethnic group | 0 (0) | 0 (0) |
| Mixed - White & Asian | 0 (0) | 1 (6.7) |
| **Stakeholder group** |  |  |
| People with lived mental health experience | 8 (80.0) | 0 |
| Family carer/friend | 2 (20.0) | 0 |
| Health and Social Care professional (statutory) | 0 | 5 (33.3) |
| Representative of a voluntary sector/charity organization | 0 | 10 (66.7) |
| **Residence/Workplace** |  |  |
| Norfolk | 10 (100) | 11 (73.3) |
| Suffolk | 0 (0) | 6 (40) |
| **Migration** |  |  |
| Moved to East Anglia within 10 years (yes) | 3 (30) | 5 (33.3) |
| Moved within the UK | 3 (30) | 4 (26.7) |
| Moved from outside of the UK | 0 (0) | 1 (6.7) |
| **Geographical settings** |  |  |
| Urban | 7 (70) | 4 (26.7) |
| Rural | 3 (30) | 9 (60) |
| Coastal | 0 (0) | 2 (13.3) |

### The Top 10 mental health research priorities

Table 5 summarises the average scores and ranking of the shortlisted research priority statements from both workshops. Participants from both workshops considered all research priority statements as important and urgent. They recommended and agreed that the tied scores should be ranked equally (See Table 5). The final Top 10 priority statements span seven domains, including youth mental health, physical–mental health integration, access to care, impacts of rural/coastal living, social and health inequalities, health promotion and prevention, and big data solutions. Seven statements scored lower than the Top 10 threshold (see Supplementary Table S3). Interestingly, none of the research priority statements in the Migration and Mental Health domain were shortlisted or voted as high priority in this study.

**Table 5:** Top 10 mental health research priorities identified by people with lived experience, members of the public and mental health professionals.

| Top 10 (Tied Ranks) | Research Priority Statement | Lived Experience & Public Score (n=10) | Professional Score (n=15) | Combined Scores | Domain |
| --- | --- | --- | --- | --- | --- |
| 1 | Integrating the work of mental health charities with NHS services to improve **access to care** and mental health outcomes of children and young people | 16 | 26 | 42 | Children and Young People's mental health |
| 1 | Improving access to **psychological therapy-style** services for children and adolescents to prevent further deterioration of their mental health | 16 | 26 | 42 | Children and Young People's mental health |
| 1 | Improving prevention efforts for suicide and self-harm in young people | 18 | 24 | 42 | Health/Social Inequalities |
| 2 | Treating mental and physical health together by expanding physical health provisions (**e.g. physical health checks, diet**) into mental healthcare to ensure 'no one is left behind' | 13 | 27 | 40 | Physical & Mental health |
| 3 | Improving access to mental health care for all age groups | 15 | 24 | 39 | Access to care |
| 3 | Addressing the barriers to accessing care and treatment for mental health difficulties in rural, remote and coastal areas | 17 | 22 | 39 | Access to care |
| 4 | Empowering patients, relatives, and carers to have a greater say about their choices regarding mental health and mental healthcare | 17 | 21 | 38 | Prevention and Promotion |
| 5 | Improving prevention efforts for suicide and self-harm in other jobs with higher risk of suicide | 14 | 22 | 36 | Health/Social Inequalities |
| 6 | Understanding which models of care increase access and support decision-making for people with mental health difficulties | 15 | 20 | 35 | Access to care |
| 6 | Increasing access to psychological/emotional support for **family carers/informal carers** of people with mental health difficulties | 16 | 19 | 35 | Access to care |
| 6 | Early intervention to prevent mental health problems in people with disabilities | 12 | 23 | 35 | Prevention and Promotion |
| 6 | Supporting parents or family members in identifying early signs of mental health difficulties in children and young people | 14 | 21 | 35 | Prevention and Promotion |
| 6 | Support for carers of older people with multiple conditions to maintain their psychological well-being | 16 | 19 | 35 | Prevention and Promotion |
| 7 | Identifying the most effective and acceptable strategies for preventing multiple physical and mental health conditions in later life | 16 | 18 | 34 | Physical & Mental health |
| 8 | Increase social and economic opportunities to enhance the mental health and **wellbeing for young people and** adults living in rural and coastal communities | 13 | 20 | 33 | Impacts of Rural/Coastal Living |
| 9 | Involving young people in making decisions about their mental health treatment | 11 | 21 | 32 | Children and Young People's mental health |
| 9 | More collaboration between health, social care, public health and voluntary sectors to **improve access** to and deliver mental health care to people in rural and coastal areas | 9 | 23 | 32 | Impacts of Rural/Coastal Living |
| 9 | Understanding how effective community-based interventions are in improving mental health in deprived areas | 10 | 22 | 32 | Health/Social Inequalities |
| 10 | Develop culturally sensitive care for racial and ethnic minoritised groups | 9 | 22 | 31 | Health/Social Inequalities |
| 10 | Improving prevention efforts for suicide and self-harm in farmers and agricultural communities | 11 | 20 | 31 | Health/Social Inequalities |
| 10 | Identify mental health policy/interventions (e.g. **support finding and retaining employment**) that could prevent poor mental health outcomes | 13 | 18 | 31 | Health/Social Inequalities |
| 10 | Using de-identified data (e.g. health, education, local authority records) to **identify risk factors** and examine mental health outcomes | 11 | 20 | 31 | Big data solutions |

Bold entries indicate phrases/words mapped from similar statements within the same domain

#### Qualitative data- group discussion in workshops

During the workshop discussions, participants reflected on how difficult it was to exclude any of the priority statements due to their importance. When discussing the scores and ranks, they highlighted that several priority statements within each domain were interconnected or similar. Consequently, they identified and mapped eleven statements onto the final Top 10 priority list (see Table 5, highlighted in bold). Our thematic analysis revealed four key themes across the 2-day workshops: including reflections, impact and aspirations, challenges, and suggestions for future directions.

### Reflections on the Workshops

Participants appreciated the opportunity to engage in meaningful discussions about mental health services and research priorities in Norfolk and Suffolk. They described the exercise as empowering, enlightening, and a valuable chance to connect with people they would not usually meet. The professional participants highlighted that the process of reviewing priorities from those with lived experience had been crucial and respectful.

Participants from the lived experience group commented that group discussions helped clarify meanings and core issues, and that listening to others’ experiences was powerful and validating. They highlighted the importance of co-production and reported feeling safe, heard, and valued.

### Challenges and Considerations

Both the professional and lived experience participants recognised and commented that navigating existing mental health services is difficult—even for professionals—highlighting the need for clearer care pathways. There was an emphasis on prevention and understanding the social, economic, and geographical factors affecting mental health. Participants also expressed concerns about the limited or lack of interventions for individuals who do not present with the severity of illness needed to meet the criteria of mental health services.

### Impact and Aspirations

Participants commented that they felt confident in the research process and reported feeling hopeful that research would lead to tangible changes in mental healthcare, including accessible, timely, consistent, and sustainable mental health care. They emphasised the importance of holistic care, early intervention, and research outcomes that influence policy and guidelines, leading to sustainable change.

### Suggestions for Future Direction

Participants offered suggestions for future research and service development, including focusing on young people and older rural populations in Norfolk and Suffolk. They shared decision-making and novel approaches tailored to local needs.

## DISCUSSION

### Main findings

In line with the JLA priority-setting framework, this study employed a mixed-methods approach to consult with and collaborate with people with lived experience of mental health difficulties, mental health experts, policymakers, and members of the public, and to identify priorities and capacity gaps for mental health research in Norfolk and Suffolk. Through the two-day workshops, mental health stakeholders in rural and coastal communities demonstrated an insight of local mental health needs and identified clear research priorities in seven topic areas, including health inequalities, impacts of rural and coastal living, children and young people’s mental health, treating physical and mental health together, improving access to appropriate care, use of big data and health promotion. These insights demonstrate that existing priorities identified for health and social care [37] are relevant and useful to rural and coastal communities.

### Methodological considerations

#### Strengths

This is one of the handful of studies available that have specifically engaged people living and working in rural and coastal communities in identifying mental health research priorities in the UK. A key strength lies in the co-production of research priorities with multiple mental health stakeholders, including patients, the public, clinicians, the voluntary sector, and policymakers, and in their guidance by a steering group and LEAG. The JLA framework and methodology allowed lived experience and healthcare professional voices to be heard, and our systematic, transparent approach aligns with other evidence-based priority research activities [39]. Our mixed-methods approach meant that we were able to capture contextual insights that shaped and refined the final research priorities through participant co-production. Demographically, the majority people of our sample were people of white British ethnic background and mostly female, these chime with the ethnic composition Norfolk and Suffolk population, where 93.9% are of white British ethnic background and 51% females [44]. Other areas of strength are the multidisciplinary and interdisciplinary partnerships, the involvement of key local and regional stakeholders from inception, and strong collaboration with other established mental health research institutions. While we observed some differences between priorities, there are substantial overlaps between patients’ and professional groups’ perspectives in mental health research priorities.

#### Limitations

Despite its strengths, our study is limited by several factors. First, we included participants aged 16 and over. Although family carers or parents of children with mental health difficulties were included, future mental health research priority-setting involving children is warranted. Second, whilst we recruited a diverse group of participants in terms of gender, ethnicity, stakeholder expertise, and rurality the small sample from coastal communities despite our targeted recruitment strategies highlights challenges in reaching participants in these areas. Proactive future research reaching out to coastal communities is needed. Third, we included participants who were fluent in English language, due to the limited study resources. Insights from non-English speakers would have strengthened our results. Future studies incorporating translation of study materials to other languages or employing interpreters are recommended. Further, most participants both in the survey and prioritisation workshops were from a white British background however, as noted above, there is a smaller proportion of people from ethnic minority backgrounds in our study catchment areas. Nonetheless, engaging minoritised ethnic people in health research could be strengthened through collaboration with community organisations that represent these groups.

#### Comparisons with other studies

Reports of mental health research priority setting exercises in the UK’s rural and coastal communities are few. The top 10 priorities identified in our study align with those in other international studies. For example, Eley and Baker (2007) engaged healthcare providers in interactive workshops to explore their perspectives on key issues affecting community health services in rural southern Queensland, Australia. Their participants identified key priorities, including mental health care, access to health services, consumer perceptions [45]. Similarly, Kshatri, Bhaumik et al. (2025) identified social determinants, mental health, the use of electronic health records, and healthcare resilience as key priority areas [46]. Additionally, Chapman, Browning et al. (2022 in a national priority setting exercise for difficult-to-treat-depression identified pharmacological, psychological as top priority treatment approaches [47]. Furthermore, the identified priorities in our study chime with the insights from our in-depth discussion with people with lived experience of mental ill-health across Norfolk and Suffolk, where participants highlighted early intervention, support for family and carers and integrated care as key priority areas [48].

### Implications and Future Research

Our approach has generated an agenda for mental health research priorities for Norfolk and Suffolk with relevance to other rural and coastal communities. Our findings demonstrate that it is feasible to employ a systematic methodology for research priority setting that includes stakeholders across health and social care systems. Our findings have implications for service planning, delivery, policymaking and future research. Children and young people’s mental health was ranked as the number 1 priority, suggesting that an emphasis on early intervention and prevention of poor outcomes in later life is key. This is particularly important in remote, rural and coastal communities where access to mental healthcare is limited; therefore, schools would be key platforms for delivering mental health programmes for young people. Integrating physical and mental healthcare was ranked the number 2 priority, highlighting the need for delivering holistic care, hence reducing poor health outcomes among people with mental health difficulties. Overall, the social and health inequalities domain had the most statements in the top 10 list, indicating consensus on reducing inequalities. The high-ranking research priority statements in this domain relate to social determinants of health and to developing interventions for specific populations, such as farming communities, those living in deprived areas, and those facing employment challenges, with a focus on cultural sensitivity. There were four priority statements relating to access to care, reflecting the challenges posed by limited resources and mental health provision across all age groups, particularly in rural and coastal communities. Prevention and health promotion were also prominent in the Top 10 list, which included four statements about family carers, parents, and relatives of people with mental health difficulties. This highlights the importance of reducing the detrimental effects that an unpaid caring role can have on the well-being of family and relatives. Whilst only one priority statement related to the use of de-identified electronic health and other administrative records for research, this reflects stakeholders’ appreciation and awareness of the value of such data sources for including people who would otherwise be underrepresented in research. Additionally, their large-scale nature allows investigation of a range of mental health outcomes independent of exposures.

## CONCLUSIONS

Until now, the lived experience voice in mental health research priority-setting from rural and coastal communities has been absent, despite the public health importance of mental health and the role of PPIE in good research practice. This study addressed this gap by (1) identifying the Top 10 actionable mental health priorities for mental health research for Norfolk and Suffolk; (2) demonstrating that large-scale stakeholder involvement is feasible in mental health research agenda setting; and (3) by showing that while there are some differences between priorities, there are substantial overlaps between patients and professional groups perspectives in mental health research priorities. These insights will shape future research and guide policymaking and resource allocation efforts to improve mental healthcare and reduce treatment gaps in rural and coastal communities.

## Data Availability

All data produced in the present study are available upon reasonable request to the authors

## FUNDING

This project is funded by the National Institute for Health and Care Research (NIHR) under its Mental Health Research Development Award Programme (Grant Reference Number NIHR207498). The views expressed are those of the author(s) and not necessarily those of the NIHR or the Department of Health and Social Care.

## ACKNOWLEDGEMENTS

The authors thank all the members of our Steering Group and Lived Experience Advisory Group for their contributions to the project.

## AUTHORS’ CONTRIBUTIONS

SO is the Chief Investigator and conceived the original study design.

Data analysis- SO, SM, JB

Writing – Original draft: SO

Writing – reviewing and editing: all authors.

All authors read and approved the final manuscript.

## CONFLICTS OF INTEREST

On behalf of all authors, the corresponding author states that there are no conflicts of interest.

## Notes

### Competing Interest Statement

The authors have declared no competing interest.

### Author Declarations

The University of East Anglia Faculty of Medicine and Health Research Ethics Committee granted ethical approval (reference ETH2324-2542).

